# Clinically relevant gene signatures provide independent prognostic information in older breast cancer patients

**DOI:** 10.1101/2023.02.14.23285920

**Authors:** Miguel Castresana-Aguirre, Annelie Johansson, Alexios Matikas, Theodoros Foukakis, Linda S. Lindström, Nicholas P. Tobin

**Affiliations:** Department of Oncology and Pathology, Karolinska Institutet and University Hospital, Stockholm, Sweden; Breast Center, Karolinska Comprehensive Cancer Center, Karolinska University Hospital, Stockholm, Sweden; Breast Cancer Now Research Unit, School of Cancer and Pharmaceutical Sciences, King’s College London, Guy’s Cancer Center, London, United Kingdom

**Author notes:** **Corresponding author:** Nicholas P. Tobin, PhD, Associate Professor, Department of Oncology and Pathology, Karolinska Institutet and University Hospital, BioClinicum, Visionsgatan 4, SE 171 64 Stockholm, Sweden.

**Keywords:** Older patients, breast cancer, gene expression signatures, prognosis

## Abstract

**Purpose:** Gene signatures have been shown to add prognostic information beyond that of routine clinico-pathological factors, however their utility in older breast cancer patients remains unclear. As such, the aim of this study was to determine if gene signatures can provide prognostic information that may aid treatment decisions for older breast cancer patients.

**Experimental design:** Research versions of the genomic grade index (GGI), 70-gene recurrence score (RS), cell cycle score (CCS), PAM50 Risk of Recurrence score - Proliferation (ROR-P), and PAM50 signatures were applied to 39 breast cancer datasets totalling 9583 patients. After filtering based on age ≥ 70 years, the presence of Estrogen Receptor (ER) and survival information availability 871 patients remained. The prognostic capacity of signatures was tested in all (n=871), ER-positive/lymph node-positive (ER+/LN+, n=335) and ER-positive/lymph node-negative (ER+/LN-, n=374) patients using Kaplan-Meier and multivariable Cox proportional hazard modeling.

**Results:** All gene signatures were statistically significant in Kaplan-Meier analysis of all and ER+/LN+ patients (Log-rank *P* < 0.001). This significance remained in multivariable analysis (Cox proportional hazards, *P* ≤ 0.05). In ER+/LN-patients the GGI, 70-gene, CCS, ROR-P, and PAM50 signatures were significant in Kaplan-Meier analysis (Log-rank *P* ≤ 0.05) but only the 70-gene, CCS, ROR-P, and PAM50 signatures remained so in multivariable analysis (Cox proportional hazards, *P* ≤ 0.05).

**Conclusions:** In general, we found that gene signatures provide prognostic information in survival analyses of all, ER+/LN+ and ER+/LN-older (≥70 years) breast cancer patients, suggesting a potential role in aiding treatment decision in older patients.

**Translational Relevance:** The utility of gene expression signatures in breast cancer patients has been most clearly demonstrated in the TAILORx, RxPONDER and MINDACT randomised clinical trials. However, few older patients (≥70 years) were included in these trials meaning that signature utility in this patient group remains unclear. As such, we performed the first comprehensive study comparing the prognostic performance of multiple clinically relevant gene expression signatures in a single older breast cancer patient cohort. We show that in general gene signatures provide independent prognostic information in All, ER+/LN+ and ER+/LN-patients who are over the age of 70 years. These results support a potential role for signatures in aiding treatment decisions in older breast cancer patients and indicate that further investigation is warranted in prospective clinical study to elucidate their treatment predictive value.

## INTRODUCTION

Human life expectancy is predicted to increase globally by 4.4 years for both men and women in the coming two decades(1). This will result in a larger population of older adults and as cancer is generally a disease of aging, it is estimated that 60% of newly diagnosed cancers in 2035 will come from adults aged 65+(2). Older cancer patients are however typically underrepresented in clinical trials(3–6) and may also be undertreated relative to younger patient populations(7,8). This implies that there is a lack of data on whether the tools used to guide treatment decisions in younger (<70 years) cancer patients are also applicable to older patient populations.

Treatment decisions in early breast cancer are based on tumour size, lymph node involvement, stage and prognostic and predictive biomarkers including the estrogen, progesterone and human epithelial growth factor 2 receptors (ER, PR and HER2) and expression of the proliferation marker Ki67(9). A patient’s age is also recommended to be taken into consideration under the provision that it should not be used as a reason to withhold specific treatments(9). This is in line with data from the Early Breast Cancer Trialists Collaborative Group (EBTCG) showing that the relative benefit from chemotherapy is independent of age(10), and international treatment guidelines for older breast cancer patients (>70 years) which state that endocrine treatment should be offered to postmenopausal women irrespective of age(11). It is important to note however that treatments should only be offered after an initial screening assessment for frailty(11).

In addition to routine pathological staging and clinical biomarkers, recent years have also seen an increase in the use of multigene signatures to aid in risk stratification of early breast cancer patients. The signature field is most mature in breast cancer with the 21-gene recurrence score (RS) and 70-gene signatures demonstrating prognostic capacity in large scale randomized clinical trials(12–14). Moreover, these signatures along with others can be used to guide treatment decisions primarily in postmenopausal early breast cancer patients with node negative or positive (1-3 nodes) invasive tumours(15). The use of gene signatures in older breast cancer patients remains controversial however as there is currently insufficient evidence to support their use(11). As such, we aimed to perform the first comprehensive comparison of the additional prognostic capacity of clinically relevant breast cancer gene signatures beyond that of routine biomarkers in a single older breast cancer cohort. Specifically, we apply the genomic grade index (GGI), 70-gene, 21-gene recurrence score (RS), cell cycle score (CCS), PAM50 Risk of Recurrence score - Proliferation (ROR-P), and PAM50 signatures to 39 open access breast cancer datasets with a combined total of 871 patients aged 70 years or older.

## PATIENTS AND METHODS

### Cohort description

The data for this study was extracted from the R package MetaGxBreast(16), a gene expression database of 39 open access breast cancer datasets with manually-curated and standardized clinical, pathological, survival, and treatment metadata for breast cancer totalling 9583 patients. After first selecting patients who were aged 70 years or older (n = 1399), samples were subsequently excluded on the basis of: lacking information on ER status (n = 62), lacking survival information (n = 323), or insufficient coverage of gene signature genes (n = 143, further described below), 871 patients remained in total (see CONSORT diagram in Figure 1). We further stratified these patients into clinically relevant subgroups of ER+/LN+ (N = 374) and ER+/LN-(N = 335) to analyse the prognostic capacity of gene signatures taking ER and lymph node statuses into account. Information regarding the number of positive lymph nodes was not available. Importantly, in this study we define older patients as those over the age of 69 years, in line with previous publications(17–21) and randomized clinical trials(22,23). In order to compare older patients to a younger postmenopausal breast cancer patient population we also selected ER+/LN-patients between 55 and 65 years of age (N = 478, labelled as “55 - 65” in subsequent analyses). This 5-year gap between 65 and 70 years was selected to have a clear separation by age and the 10 year interval was chosen in order to obtain a large enough cohort for comparison.

**Figure 1:**
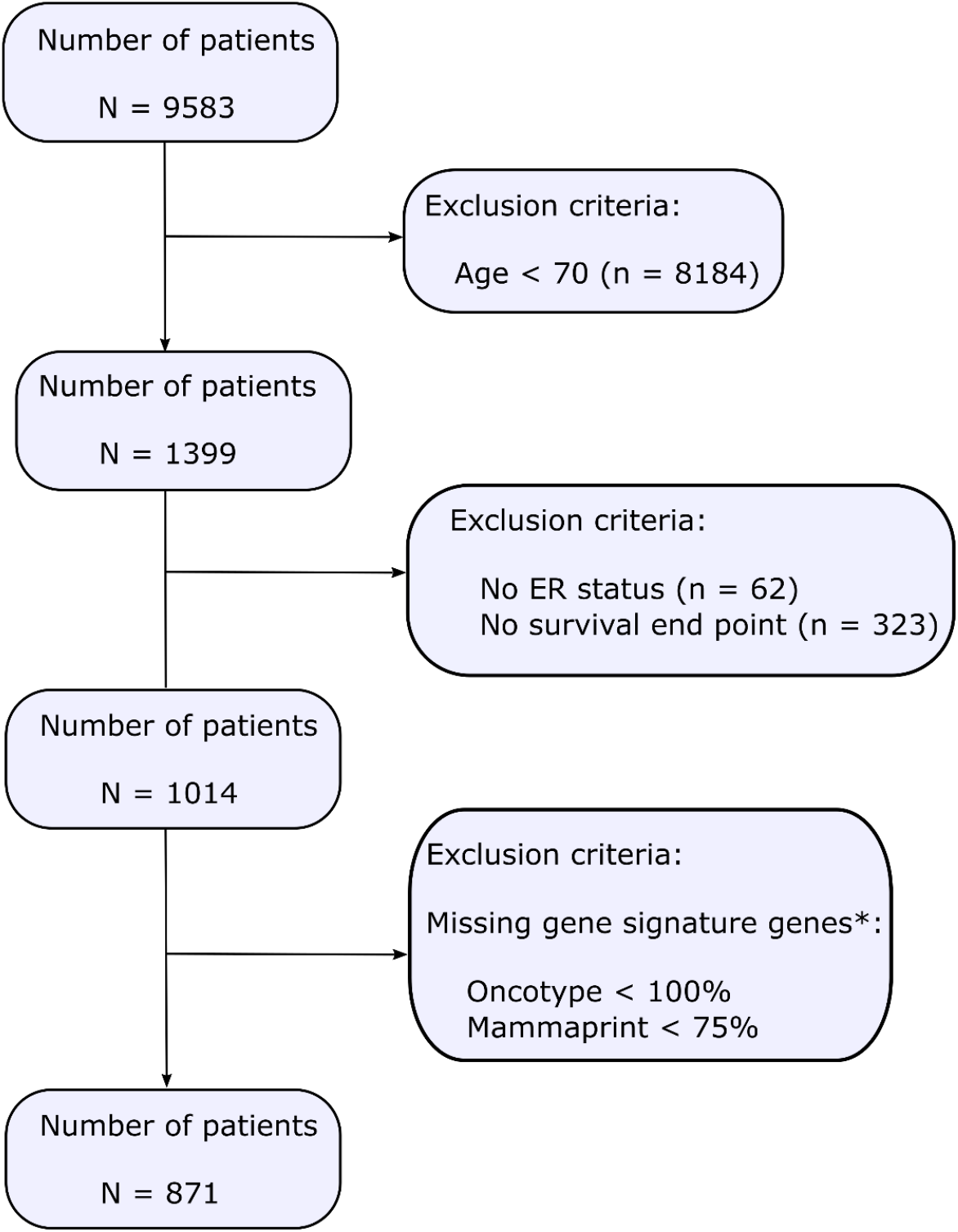
Consort diagram of older breast cancer patient selection from the 39 datasets in the MetaGxBreast database. Patients were excluded owing to lacking information on ER, being under the age of 70 years old, lacking survival information or having insufficient coverage of gene signature genes. * See methods for a full description.

**Figure 2:**
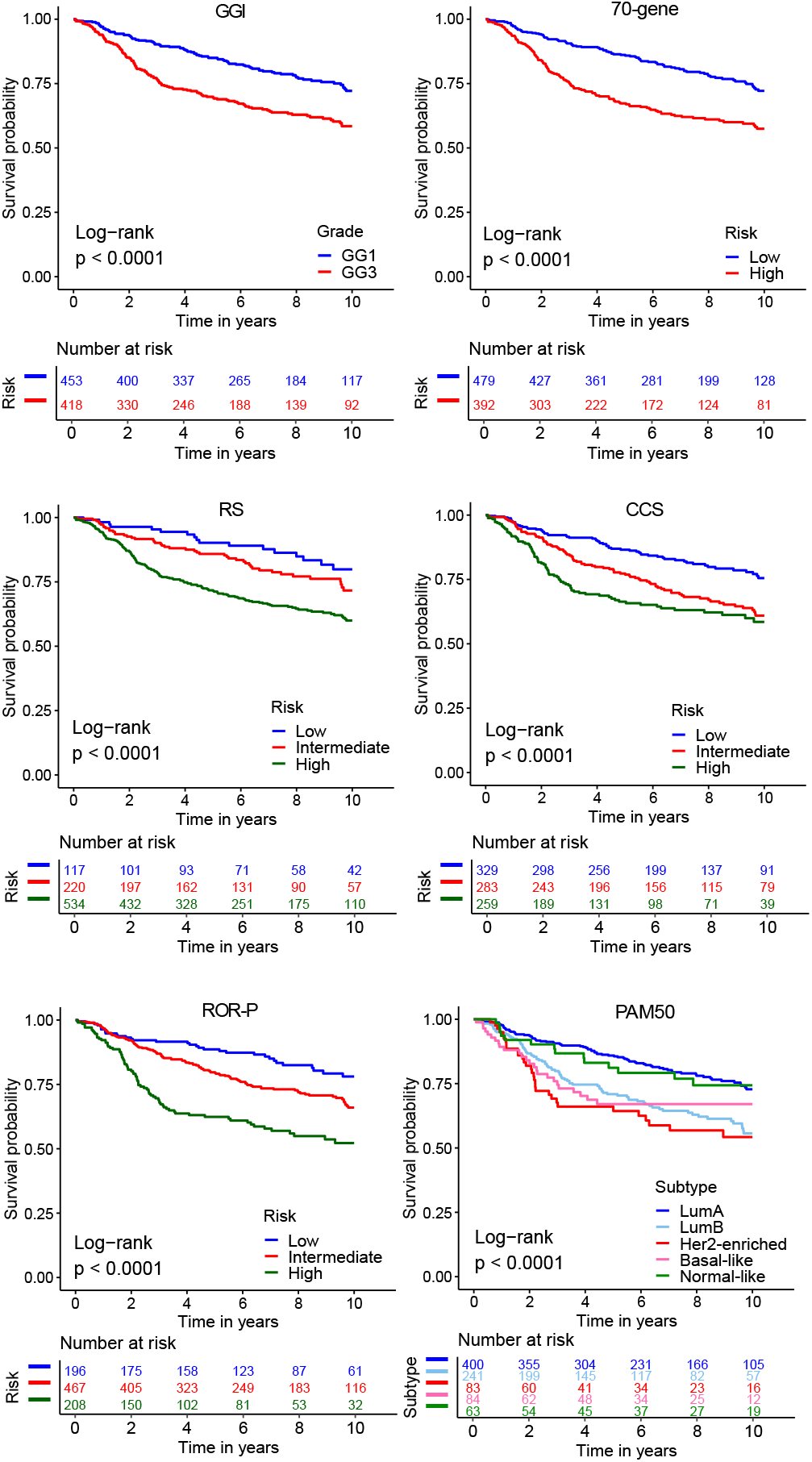
Kaplan-Meier analysis of gene expression signatures in All patients of the older cohort. (a) Genomic Grade Index (GGI) (b) 70-gene (c) Recurrence score (RS) (d) Cell-cycle (CCS) (e) PAM50 Risk of Recurrence score - Proliferation (ROR-P) (f) Prediction Analysis of Microarray 50 (PAM50).

### Gene expression signatures

Research versions of the Genomic Grade Index (GGI)(24), 70 gene(25), 21-gene recurrence score (RS)(26), Cell Cycle Score (CCS)(27), PAM50 Risk of Recurrence score - Proliferation (ROR-P), and Prediction Analysis of Microarray 50 (PAM50)(28), were applied on an individual dataset basis to each of the 39 MetaGxBreast studies as described in the original publications. Signature classifications were subsequently pooled with clinical data for statistical analysis. For GGI, tumour grade was not available for all patients (missing in 38%), as such, we used a variation of the tool to compute the tumour grade(29) and then ran the conventional research version of GGI. The original RS signature cutoffs (Low < 18, Intermediate 18 - 31, High > 31) were used throughout the study, however the updated cutoffs from the TAILORx clinical trial (Low < 10, Intermediate 11 - 25, High > 26) were also assessed for the sake of completeness, as specified in the results section. PAM50 and ROR-P work accurately if the dataset is ER status-balanced(30) and therefore we used Monte Carlo sampling to have equal proportions of ER+ and ER-patients. We sampled 100 subsets to compute the median of expression of the probes for centering the data(31). The gene signatures were chosen owing to their relevance in randomized clinical trials such as MINDACT(13), ASTER70(22), TAILORX(32) and also in real world evaluation(33).

### Gene expression and gene mapping

Gene expression data extracted from MetaGxBreast comes pre-processed and normalized, a detailed description of this can be found in the original publication(16). Probe to gene mapping was achieved by merging annotation sources from MetaGxBreast, supplementary files from the original signature publications and Bioconductor 3.15 in R. Probes mapping to the same gene were combined by averaging their expression values. MetaGxBreast combines datasets from different gene expression array platforms, the majority of which are Affymetrix. This means that all genes are not found in all datasets. The 70-gene signature is derived on an Agilent array platform, and approximately 75% of the signature’s genes are mappable to the Affymetrix platform. For this reason, we excluded datasets if less than 75% of the 70-gene signature probes were present and similarly if any non-reference genes for the RS were absent. Consequently, the average gene coverage for GGI was 100%, 98% for PAM50, 89% for the 70-gene signature, 100% for RS, and 97% for CCS.

### Statistical analysis

Kaplan–Meier and multivariable Cox proportional hazard analyses were used to assess older patient survival in the context of gene expression signature subgroups. The latter was adjusted for ER status, lymph node status, tumour grade, tumour size and whether the patients had received hormonal therapy or not. We did not adjust for treatment with chemotherapy as few patients received it (N = 50). These methodologies were applied to All patients as well the subgroups of ER+/LN- and ER+/LN+ patients. Subgroups were adjusted for tumour size, grade, and hormonal therapy only. The clinical endpoint used was Recurrence Free Survival (RFS) defined as the time from date of curative surgery to the time recurrence (distant metastatic events and loco-regional recurrences). RFS was censored at 10 years and the median follow-up time was 6.2 years. MetaGxBreast does not provide RFS data for METABRIC, so this information was instead extracted from the supplementary data of Rueda et al.(34). The likelihood ratio (LR) and the concordance index (c-index) were computed using univariable models as a measure of signature prognostic capacity. For determination of the additional prognostic capacity of signatures beyond clinico-pathological markers we calculated the delta likelihood ratio (ΔLR) by comparing the LR of a multivariable model that included the adjustment variables noted above with and without the gene expression signature. To assess if there was any statistical difference in the classification of patients by gene signatures between younger postmenopausal patients (between 55 and 65 years of age) and older patients, we performed Chi-squared tests. Tests used are indicated in table legends. All statistical tests were two-sided and a significance level ⍺ of 5 % was used. All statistical analyses were performed using R statistical software version 4.1.2.

## Data availability

The data used in this study are publicly available on MetaGxBreast package in R (https://bioconductor.org/packages/release/data/experiment/html/MetaGxBreast.html). R-code to reproduce the results of this study are publicly available at https://bitbucket.org/tobingroup/elderly

## RESULTS

### Cohort clinico-pathological characteristics and gene signature distribution

In order to assess the prognostic capacity of gene expression signatures in older (≥ 70 years) breast cancer patients, we applied research versions of the GGI, 70 gene, RS, CCS, ROR-P, and PAM50 signatures to expression array data from 39 open access breast cancer datasets individually. Signature classifications and clinico-pathological variables were then merged into a single dataset. After limiting the cohort to an older population only and filtering on the basis of exclusion criteria (see methods), 871 patients remained (CONSORT diagram, Figure 1). The median age of these patients was 75 years old (range 70 to 96) and as expected, a decrease in age frequency is readily apparent as patients tend towards the upper age range (Supplementary Figure 1). Clinico-pathological characteristics for these patients are shown in Table 1. The majority of patients were ER-positive (87%), PR-positive (47%) and HER2-negative (65%, Table 1, PR and HER2 status unknown in 23 and 27% of patients, respectively). Half of all patients were negative for lymph node metastases and the majority of tumours were of larger size (69% ≥2 cm) and intermediate or high grade (81%). 57% of patients received hormonal therapy but few received chemotherapy (<6%), as anticipated given that many of the datasets used in this study are from cohorts assembled before the 2000s where treatments were less standardized. The number of patients categorized into subgroups on the basis of gene expression signatures is shown in Supplementary Table 1. Binary signatures demonstrated an approximate even split into grouping patients into low and high-risk groups (see GGI and 70-gene, Supplementary Table 1). Signatures with three levels (RS, CCS and ROR-P) classified the majority of patients into intermediate and high-risk groups and PAM50 classified 74% of patients into luminal A or B subtypes, in line with the high level of ER-positive patients in the cohort.

**Table 1:**
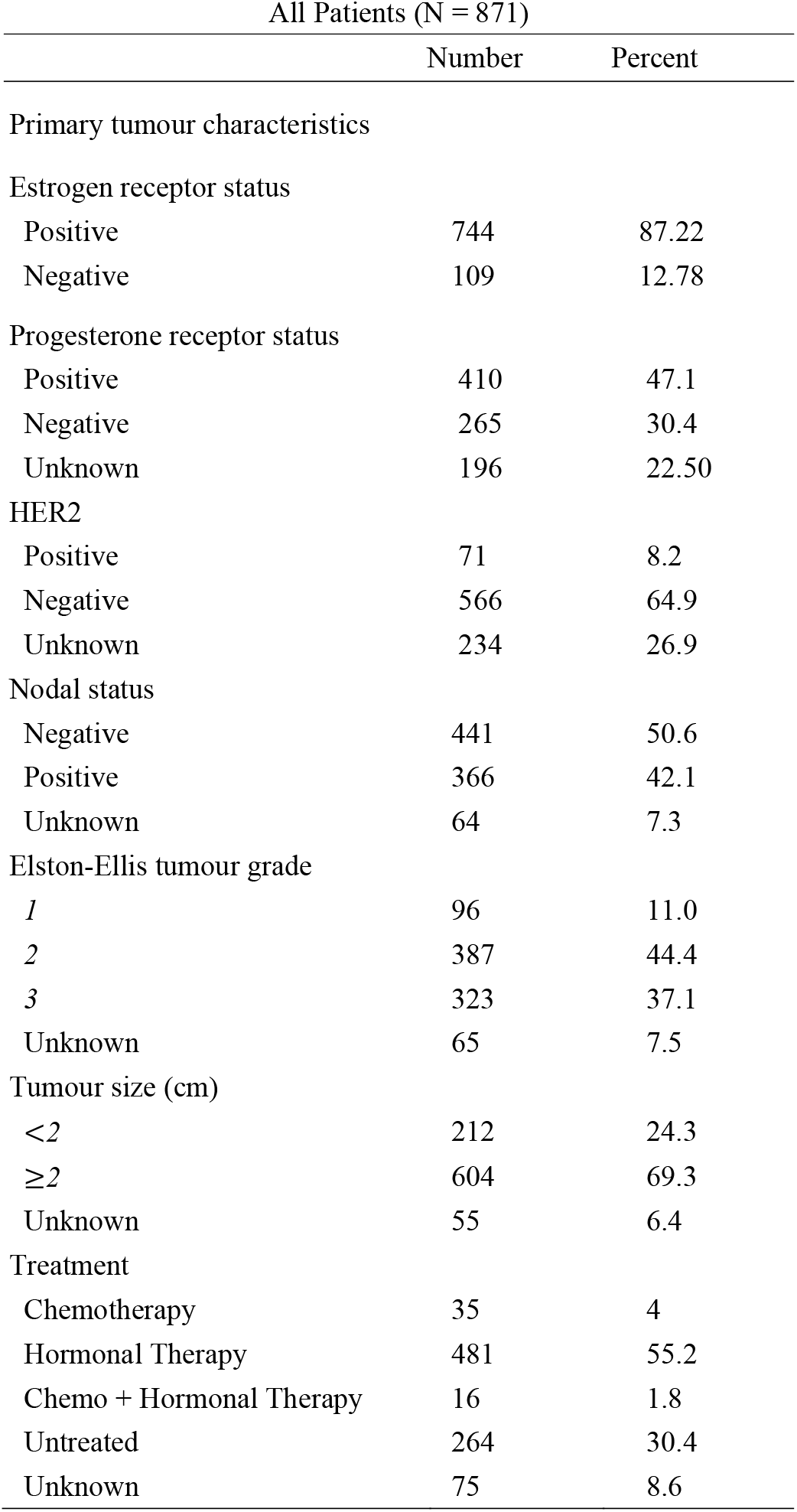
Clinico-pathological characteristics of the older breast cancer cohort.

### Gene signatures provide independent prognostic information for older breast cancer patients

We next assessed the prognostic capacity of the GGI, 70-gene, RS, CCS, ROR-P, and PAM50 gene signatures in All (*N* = 871), ER+/LN+ (*N* = 335), and ER+/LN-patients (*N* = 374) using Kaplan-Meier analysis. All signatures provided prognostic information in All and ER+/LN+ patients (log-rank test, *P* < 0.05, Figure 1 and Supplementary Figure 2). In the ER+/LN-subgroup, the GGI, 70-gene, CCS, ROR-P and PAM50 signatures show statistical significance (log-rank test, *P* < 0.05, Supplementary Figure 3) with a non-significant trend observed for RS(log-rank test, *P* = 0.068, Supplementary Figure 3). In general, statistical significance remained in multivariable Cox proportional hazards analyses for all signatures in All and ER+/LN+ subgroups after adjusting for tumour size, tumour grade, ER status, lymph node status, and whether the patient received hormonal therapy or not (Cox proportional hazards modeling, *P* < 0.05 vs. signature reference group. Table 2). In ER+/LN-patients however, only the 70-gene, CCS, ROR-P and PAM50 signatures remained statistically significant in the same analysis (Table 2). Of note, we also analyzed ER+/LN-/HER2-patients and we observed similar trends (Supplementary Table 2) as for ER+/LN-patients but only the 70-gene signature and PAM50 showed statistically significant independent prognostic information (*P* = 0.02 for 70-gene high vs. low risk and *P* = 0.03 for PAM50 Luminal B vs Luminal A). We also applied Likelihood ratio statistics to assess the additional prognostic capacity of signatures beyond routine clinico-pathological markers. The largest ΔLRs were found for the RS, CCS and ROR-P signatures in All (ΔLR = 14.11, 16.97, and 22.42) and ER+/LN+ (ΔLR = 13.6, 13.2 and 13.88) patients, respectively (LR-test, *P* < 0.01, Supplementary Table 3). Lower ΔLRs were found for signatures in ER+/LN-patients (ΔLR = 5.88, 6.47 and 5.49 for the 70 gene, CCS and ROR-P signatures respectively, LR-test, *P* < 0.05, Supplementary Table 3).

**Table 2:**
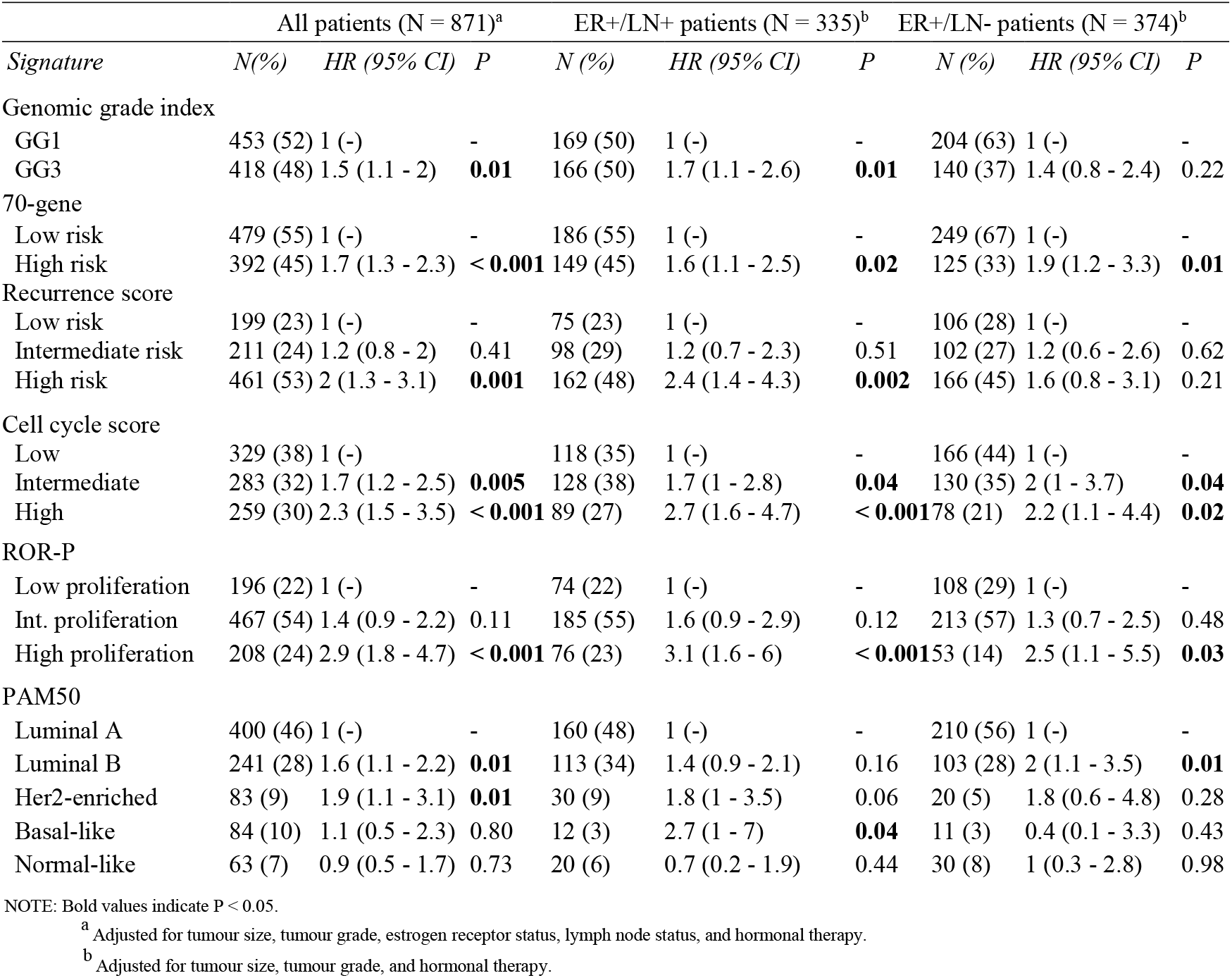
Multivariable analysis for gene signatures in All, ER+/LN+ and ER+/LN-older patients.

### Differences in signature risk stratification in comparison to a younger postmenopausal patient cohort

The clinical utility of gene signatures is most frequently discussed in relation to ER+/LN-postmenopausal breast cancer patients, however in our analysis all signatures showed reduced prognostic capacity (ΔLR) in this patient subgroup. To understand if there are differences in signature prognostic capacity between an older ER+/LN-patient cohort and a younger postmenopausal ER+/LN-cohort, we ran the same analyses but this time selecting ER+/LN-patients between 55 and 65 years old (*N* = 478). Patient characteristics for this subgroup are shown in Supplementary Table 4. We found that all gene signatures except PAM50 provided independent prognostic information in ER+/LN-patients aged 55-65 years old (Supplementary Table 5) and that the additional prognostic capacity of signatures beyond routine clinico-pathological markers were in general higher in this younger subgroup relative to older (≥ 70 years) ER+/LN-breast cancer patients (55-65 vs >70, ΔLR; GGI: 6.23 vs 1.43; 70-gene: 10.11 vs 5.88; RS: 6.59 vs 2.14; CCS: 8.93 vs 6.47; ROR-P: 12.25 vs 5.49; PAM50: 6.90 vs 8.37, Supplementary Tables 3 and 5). Of note, we also tested the TAILORx RS cutoffs in the same subgroups (55-65 and >70 ER+/LN-patients) but found it to perform worse than the original RS cutoffs – it did not provide independent prognostic information in either subgroup (data not shown).

To assess whether the generally higher LRs in 55-65 year olds could be owing to a difference in signature risk stratification (assignment of tumours into signature subgroups) we also compared signature subgroup composition between older and younger ER+/LN-patients. We found that only the ROR-P signature showed a statistically significant difference in risk stratification between these two groups (Chi-square test, *P* = 0.003, Supplementary Table 6). This suggests that the difference in signature prognostic capacity between older and younger patients is unlikely to be related to signature stratification and may point to a need for re-optimization of signatures and their prognostic cutoffs in older breast cancer populations.

## DISCUSSION

In this study we assessed the prognostic capacity of six gene expressions signatures in a cohort of 871 older (≥70 years) breast cancer patients. We found that all gene signatures provided independent prognostic information in All and ER+/LN+ patients. In ER+/LN-patients the 70-gene, CCS, ROR-P and PAM50 signatures remained statistically significant after adjusting for routine clinico-pathological variables. LR statistics showed lower additional prognostic capacity of signatures beyond these routine variables in ER+/LN-patients relative to All and ER+/LN+ patients. Further comparison of older ER+/LN-patients to a postmenopausal ER+/LN-cohort of younger age (55-65) showed higher signature ΔLR values in the younger cohort despite similar signature subgroup stratification in both groups.

To our knowledge, this is the first comprehensive study comparing the prognostic performance of multiple clinically relevant gene expression signatures in a single older breast cancer patient cohort. Previous or ongoing studies have however focused on the prognostic and treatment predictive capacity of individual signatures in older breast cancer patients. Similarly, to our findings, the ASTER 70s randomized phase III clinical trial also demonstrated that GGI is prognostic in older (≥70 years) breast cancer patients(35). The trial did not however find a statistically significant overall survival benefit with the addition of chemotherapy to endocrine therapy after surgery in ER+/HER2-patients whose tumours were classified as GGI high (GG3)(35). A recent study of the 70-gene signature in 89 older patients showed no statistical difference from the MINDACT clinical trial implying that this signature could also be applied to older breast cancer patients(36). Furthermore, in a study with 418 older patients above 70, Noordhoek *et al*. found that patients with high clinical risk based on the St. Gallen risk classification, but classified as ultra-low risk by the 70-gene signature, had excellent prognosis(37). Regarding RS, Iles *et al*. showed a decline in its usage with increasing age and a higher prevalence of low-risk classifications in patients above 70 years old(38). ER+/LN-/HER2-patients derive low or no benefit from chemotherapy if a tumour is classified as RS low(32) and in line with this, Barni *et al*. found a 38% relative reduction of chemotherapy usage in 230 older patients on the basis of RS(39). Whilst the above studies focus on de-escalation, age alone should not be a contraindication for chemotherapy usage(40–42). This implies that running gene signatures on older breast cancer patients could also identify those who would benefit from chemotherapy treatment but may not in the past have been treated owing to advanced age. RS has proved useful for escalation of chemotherapy in older patients, with a treatment decision change of 18.6% mainly from no-chemotherapy to chemotherapy in a cohort of 237 patients(43). Interestingly, in our study gene expression signatures classified between 25 and 53% of all tumours into high-risk groups however, only 6% of the patients received chemotherapy, none of whom were ER+/LN-.

Breast cancer clinical oncology guidelines support the use of the 70-gene (commercially MammaPrint) and RS (OncotypeDx) signatures to guide endocrine or chemotherapy treatment decisions in postmenopausal ER+/LN-/HER2-patients (or with 1-3 positive lymph nodes) and PAM50 (Prosigna) in postmenopausal ER+/LN-/HER2-patients(9,15). In our study, the 70-gene and PAM50 signatures provided independent prognostic information in ER+/LN-older patients (*N* = 374) and showed similar trends in ER+/LN-/HER2-patients (*P* ≤ 0.05, *N* = 222). However, we did note that the additional prognostic capacity of signatures beyond routine clinico-pathological markers was reduced in our older ER+/LN-patient cohort relative to those aged 55-65. The reason for this is not readily apparent but one potential concern is that the cutoffs for signature classification into prognostic groups were optimized on a younger patient population and may need to be re-optimised/changed for application to an older cohort. Related to this, Jezequel *et al*. noted a difference in the proportions of patients classified into good/poor prognosis groups by GGI, 70-gene and RS when comparing the age groups ≤40, 40-70, and ≥70(44). Similarly, Kruijf *et al*. found differences in PAM50 subtype proportions and weaker signature prognostic capacity in older patients (≥65) when compared to a younger (<65) patient population(45). In addition, an increase in the proportion of tumours classified as luminal subtypes and a decrease in basal-like subtypes has also been previously reported in older patients(44–46). Taken together, these studies imply that biomarkers and cutpoints used in younger postmenopausal breast cancer population might not be directly transferable to older patients without modification. In the current study only the ROR-P signatures showed a statistically significant difference in risk stratification when comparing older vs. younger postmenopausal patients (Supplementary Table 6), but non-significant trends were noted for GGI and PAM50. As such these differences are unlikely to explain the reduced additional prognostic capacity of signatures in ER+/LN-older patients and this reduction could be owing to a difference in the biology of tumours from older populations. This is also supported by one study showing that luminal B tumours from patients over the age of 70 years were less aggressive than those from younger age groups and that this was related to differences in pathways for iron metabolism, mitochondrial oxidative phosphorylation and reactive stroma(44).

The strengths of our study are as follows. First, we provide a comprehensive analysis of the prognostic capacity of six gene expression signatures in an older breast cancer patient cohort with a median age of 75.4 years old (52% of patients were over the age of 75). This is notable as no patients above the age of 75 were included in the TAILORx or MINDACT clinical trials and only 12% of patients were above 70 years old in the RxPONDER trial(14), despite 30% of the breast cancer diagnosis occurring in patients above 70(47). Second, we assess the additional prognostic capacity of these signatures beyond routine clinico-pathological biomarkers - something that is currently lacking in published literature. There are also some limitations to this study. First, no clear definition of what constitutes a patient as “older” is routinely applied in a clinical setting. Even though we defined this as patients ≥70 years of age, the usage of chronological age may ignore the diverse ways that time affects individuals. Since cancer is a disease of aging(48), a better definition of older could be obtained using the biological age which takes multiple biological and physiological developmental factors including genetics, lifestyle, diet and comorbidities(49). Related to this, a second limitation is that we did not adjust our analyses for patient frailty which is known to negatively impact prognosis(50), owing to no data on this being available for this metric. Third, clinical survival endpoints were not identical across the 39 independent datasets we assessed; therefore, we combined two different end points (RFS and DMFS) into a single survival metric. Of note, we chose not to use an overall survival (OS) endpoint owing to potential competing causes of death. Fourth, we relied on the research versions of gene-expression signatures in place of their commercial implementations, fifth, we lack patient numbers to assess the treatment predictive value of these signature and sixth the CCS risk stratification cutoffs were derived from the METABRIC dataset which is included in our study. Whilst we have never optimized the CCS cutoffs for prognostic capacity, there is still the potential for an overfit bias that could possibly impact our results for this signature only.

In conclusion, we show that gene expression signatures provide independent prognostic information in All, ER+/LN+ and ER+/LN-patients who are over the age of 70 years, supporting the rationale of the ASTER70s clinical trial. These results suggest a potential role for gene expression signatures in aiding treatment decisions in older breast cancer patients and indicate that further investigation is warranted in prospective clinical study to elucidate their treatment predictive value.

## Data Availability

https://bioconductor.org/packages/release/data/experiment/html/MetaGxBreast.html

## REFERENCES

1. Foreman KJ, Marquez N, Dolgert A, Fukutaki K, Fullman N, McGaughey M, et al. Forecasting life expectancy, years of life lost, and all-cause and cause-specific mortality for 250 causes of death: reference and alternative scenarios for 2016-40 for 195 countries and territories. Lancet. 2018;392:2052–90.

2. Pilleron S, Sarfati D, Janssen-Heijnen M, Vignat J, Ferlay J, Bray F, et al. Global cancer incidence in older adults, 2012 and 2035: A population-based study. Int J Cancer. 2019;144:49–58.

3. Townsley CA, Selby R, Siu LL. Systematic review of barriers to the recruitment of older patients with cancer onto clinical trials. J Clin Oncol. 2005;23:3112–24.

4. Hutchins LF, Unger JM, Crowley JJ, Coltman CA Jr, Albain KS. Underrepresentation of patients 65 years of age or older in cancer-treatment trials. N Engl J Med. 1999;341:2061–7.

5. Lewis JH, Kilgore ML, Goldman DP, Trimble EL, Kaplan R, Montello MJ, et al. Participation of patients 65 years of age or older in cancer clinical trials. J Clin Oncol. 2003;21:1383–9.

6. Talarico L, Chen G, Pazdur R. Enrollment of elderly patients in clinical trials for cancer drug registration: a 7-year experience by the US Food and Drug Administration. J Clin Oncol. 2004;22:4626–31.

7. Bouchardy C, Rapiti E, Fioretta G, Laissue P, Neyroud-Caspar I, Schäfer P, et al. Undertreatment strongly decreases prognosis of breast cancer in elderly women. J Clin Oncol. 2003;21:3580–7.

8. Noon AP, Albertsen PC, Thomas F, Rosario DJ, Catto JWF. Competing mortality in patients diagnosed with bladder cancer: evidence of undertreatment in the elderly and female patients. Br J Cancer. 2013;108:1534–40.

9. Cardoso F, Kyriakides S, Ohno S, Penault-Llorca F, Poortmans P, Rubio IT, et al. Early breast cancer: ESMO Clinical Practice Guidelines for diagnosis, treatment and follow-up†. Ann Oncol. 2019;30:1194–220.

10. Early Breast Cancer Trialists’ Collaborative Group (EBCTCG), Peto R, Davies C, Godwin J, Gray R, Pan HC, et al. Comparisons between different polychemotherapy regimens for early breast cancer: meta-analyses of long-term outcome among 100,000 women in 123 randomised trials. Lancet. 2012;379:432–44.

11. Biganzoli L, Battisti NML, Wildiers H, McCartney A, Colloca G, Kunkler IH, et al. Updated recommendations regarding the management of older patients with breast cancer: a joint paper from the European Society of Breast Cancer Specialists (EUSOMA) and the International Society of Geriatric Oncology (SIOG). Lancet Oncol. 2021;22:e327–40.

12. Sparano JA, Gray RJ, Ravdin PM, Makower DF, Pritchard KI, Albain KS, et al. Clinical and Genomic Risk to Guide the Use of Adjuvant Therapy for Breast Cancer. N Engl J Med. 2019;380:2395–405.

13. Cardoso F, van’t Veer LJ, Bogaerts J, Slaets L, Viale G, Delaloge S, et al. 70-Gene Signature as an Aid to Treatment Decisions in Early-Stage Breast Cancer. N Engl J Med. 2016;375:717–29.

14. Kalinsky K, Barlow WE, Gralow JR, Meric-Bernstam F, Albain KS, Hayes DF, et al. 21-Gene Assay to Inform Chemotherapy Benefit in Node-Positive Breast Cancer. N Engl J Med. 2021;385:2336–47.

15. Andre F, Ismaila N, Allison KH, Barlow WE, Collyar DE, Damodaran S, et al. Biomarkers for Adjuvant Endocrine and Chemotherapy in Early-Stage Breast Cancer: ASCO Guideline Update. J Clin Oncol. 2022;40:1816–37.

16. Gendoo DMA, Zon M, Sandhu V, Manem VSK, Ratanasirigulchai N, Chen GM, et al. MetaGxData: Clinically Annotated Breast, Ovarian and Pancreatic Cancer Datasets and their Use in Generating a Multi-Cancer Gene Signature. Sci Rep. 2019;9:8770.

17. Derks MGM, Bastiaannet E, Kiderlen M, Hilling DE, Boelens PG, Walsh PM, et al. Variation in treatment and survival of older patients with non-metastatic breast cancer in five European countries: a population-based cohort study from the EURECCA Breast Cancer Group. Br J Cancer. 2018;119:121–9.

18. Wildiers H, de Glas NA. Anticancer drugs are not well tolerated in all older patients with cancer. The Lancet Healthy Longevity. Elsevier; 2020;1:e43–7.

19. Hotta K, Ueoka H, Kiura K, Tabata M, Tanimoto M. An overview of 48 elderly-specific clinical trials of systemic chemotherapy for advanced non-small cell lung cancer. Lung Cancer. 2004;46:61–76.

20. Tesarova P. Breast cancer in the elderly-Should it be treated differently? Rep Pract Oncol Radiother. 2012;18:26–33.

21. Sanabria A, Carvalho AL, Vartanian JG, Magrin J, Ikeda MK, Kowalski LP. Comorbidity Is a Prognostic Factor in Elderly Patients with Head and Neck Cancer. Ann Surg Oncol. 2007;14:1449–57.

22. Brain E, Girre V, Rollot F, Bonnetain F, Debled M, Lacroix M, et al. ASTER 70s: Benefit of adjuvant chemotherapy for estrogen receptor-positive HER2-negative breast cancer in women over 70 according to genomic grade—A French GERICO/UCBG UNICANCER multicenter phase III trial. J Clin Orthod. Wolters Kluwer; 2012;30:TPS667–TPS667.

23. Hughes KS, Schnaper LA, Berry D, Cirrincione C, McCormick B, Shank B, et al. Lumpectomy plus tamoxifen with or without irradiation in women 70 years of age or older with early breast cancer. N Engl J Med. 2004;351:971–7.

24. Sotiriou C, Wirapati P, Loi S, Harris A, Fox S, Smeds J, et al. Gene expression profiling in breast cancer: understanding the molecular basis of histologic grade to improve prognosis. J Natl Cancer Inst. 2006;98:262–72.

25. van ‘t Veer LJ, Dai H, van de Vijver MJ, He YD, Hart AAM, Mao M, et al. Gene expression profiling predicts clinical outcome of breast cancer. Nature. 2002;415:530–6.

26. Paik S, Shak S, Tang G, Kim C, Baker J, Cronin M, et al. A multigene assay to predict recurrence of tamoxifen-treated, node-negative breast cancer. N Engl J Med. 2004;351:2817– 26.

27. Tobin NP, Lundberg A, Lindström LS, Harrell JC, Foukakis T, Carlsson L, et al. PAM50 Provides Prognostic Information When Applied to the Lymph Node Metastases of Advanced Breast Cancer Patients. Clin Cancer Res. 2017;23:7225–31.

28. Parker JS, Mullins M, Cheang MCU, Leung S, Voduc D, Vickery T, et al. Supervised risk predictor of breast cancer based on intrinsic subtypes. J Clin Oncol. 2009;27:1160–7.

29. Wennmalm K, Bergh J. A simple method for assigning genomic grade to individual breast tumours. BMC Cancer. 2011;11:306.

30. Raj-Kumar P-K, Liu J, Hooke JA, Kovatich AJ, Kvecher L, Shriver CD, et al. PCA-PAM50 improves consistency between breast cancer intrinsic and clinical subtyping reclassifying a subset of luminal A tumors as luminal B. Sci Rep. 2019;9:7956.

31. Johansson A, Yu NY, Iftimi A, Tobin NP, van ‘t Veer L, Nordenskjöld B, et al. Clinical and molecular characteristics of estrogen receptor-positive ultralow risk breast cancer tumors identified by the 70-gene signature. Int J Cancer. 2022;150:2072–82.

32. Sparano JA, Gray RJ, Makower DF, Pritchard KI, Albain KS, Hayes DF, et al. Adjuvant Chemotherapy Guided by a 21-Gene Expression Assay in Breast Cancer. N Engl J Med. 2018;379:111–21.

33. Kjällquist U, Acs B, Margolin S, Karlsson E, Kessler LE, Garcia Hernandez S, et al. Real World Evaluation of the Prosigna/PAM50 Test in a Node-Negative Postmenopausal Swedish Population: A Multicenter Study. Cancers [Internet]. 2022;14. Available from: http://dx.doi.org/10.3390/cancers14112615

34. Rueda OM, Sammut S-J, Seoane JA, Chin S-F, Caswell-Jin JL, Callari M, et al. Dynamics of breast-cancer relapse reveal late-recurring ER-positive genomic subgroups. Nature. Nature Publishing Group; 2019;567:399–404.

35. Brain E, Viansone AA, Bourbouloux E, Rigal O, Ferrero J-M, Kirscher S, et al. Final results from a phase III randomized clinical trial of adjuvant endocrine therapy ± chemotherapy in women ≥ 70 years old with ER HER2-breast cancer and a high genomic grade index: The Unicancer ASTER 70s trial [Internet]. Journal of Clinical Oncology. 2022. page 500–500. Available from: http://dx.doi.org/10.1200/jco.2022.40.16_suppl.500

36. Mansani F, Celinski V, Freitas-Junior R. De-escalation of chemotherapy in elderly women using a 70-gene platform: Comparison of the MINDACT study with a real-world study in the Brazilian population (AGEMA-BRA). J Clin Orthod. Wolters Kluwer; 2022;40:e12568–e12568.

37. Noordhoek I, Bastiaannet E, de Glas NA, Scheepens J, Esserman LJ, Wesseling J, et al. Validation of the 70-gene signature test (MammaPrint) to identify patients with breast cancer aged ≥ 70 years with ultralow risk of distant recurrence: A population-based cohort study. J Geriatr Oncol [Internet]. 2022; Available from: http://dx.doi.org/10.1016/j.jgo.2022.07.006

38. Iles K, Roberson ML, Spanheimer P, Gallagher K, Ollila DW, Strassle PD, et al. The impact of age and nodal status on variations in oncotype DX testing and adjuvant treatment. npj Breast Cancer. Nature Publishing Group; 2022;8:1–8.

39. Barni S, Cognetti F, Petrelli F. Is the oncotype DX test useful in elderly breast cancer patients: a subgroup analysis of real-life Italian PONDx study. Breast Cancer Res Treat. 2022;191:477–80.

40. Brain EGC, Mertens C, Girre V, Rousseau F, Blot E, Abadie S, et al. Impact of liposomal doxorubicin-based adjuvant chemotherapy on autonomy in women over 70 with hormone-receptor-negative breast carcinoma: A French Geriatric Oncology Group (GERICO) phase II multicentre trial. Crit Rev Oncol Hematol. 2011;80:160–70.

41. Freyer G, Campone M, Peron J, Facchini T, Terret C, Berdah J-F, et al. Adjuvant docetaxel/cyclophosphamide in breast cancer patients over the age of 70: results of an observational study. Crit Rev Oncol Hematol. 2011;80:466–73.

42. Takabatake D, Taira N, Hara F, Sien T, Kiyoto S, Takashima S, et al. Feasibility Study of Docetaxel with Cyclophosphamide as Adjuvant Chemotherapy for Japanese Breast Cancer Patients. Jpn J Clin Oncol. Oxford Academic; 2009;39:478–83.

43. Yu J, Wu J, Huang O, He J, Li Z, Chen W, et al. Do 21-Gene Recurrence Score Influence Chemotherapy Decisions in T1bN0 Breast Cancer Patients? Front Oncol. 2020;10:708.

44. Jézéquel P, Sharif Z, Lasla H, Gouraud W, Guérin-Charbonnel C, Campion L, et al. Gene-expression signature functional annotation of breast cancer tumours in function of age. BMC Med Genomics. 2015;8:80.

45. de Kruijf EM, Bastiaannet E, Rubertá F, de Craen AJM, Kuppen PJK, Smit Vthbm, et al. Comparison of frequencies and prognostic effect of molecular subtypes between young and elderly breast cancer patients. Mol Oncol. 2014;8:1014–25.

46. Jenkins EO, Deal AM, Anders CK, Prat A, Perou CM, Carey LA, et al. Age-specific changes in intrinsic breast cancer subtypes: a focus on older women. Oncologist. 2014;19:1076–83.

47. Gosain R, Pollock Y, Jain D. Age-related Disparity: Breast Cancer in the Elderly. Curr Oncol Rep. 2016;18:69.

48. Berben L, Floris G, Wildiers H, Hatse S. Cancer and Aging: Two Tightly Interconnected Biological Processes. Cancers [Internet]. 2021;13. Available from: http://dx.doi.org/10.3390/cancers13061400

49. Diebel LWM, Rockwood K. Determination of Biological Age: Geriatric Assessment vs Biological Biomarkers. Curr Oncol Rep. 2021;23:104.

50. Hong C-C, Ambrosone CB, Goodwin PJ. Comorbidities and Their Management: Potential Impact on Breast Cancer Outcomes. Adv Exp Med Biol. 2015;862:155–75.

