## Supplementary Tables for "Clinically relevant gene signatures provide independent prognostic information in older breast cancer patients"

Supplementary table 1: Gene signature risk stratification of elderly patients.

| Signature | Patients ≥ 70 (N = 871) |
|---|---|
| GGI | |
| GG1 | 453 (52) |
| GG3 | 418 (48) |
| 70-gene | |
| Low risk | 479 (55) |
| High risk | 392 (45) |
| Recurrence score | |
| Low risk | 199 (23) |
| Intermediate risk | 211 (24) |
| High risk | 461 (53) |
| Cell cycle score | |
| Low | 329 (38) |
| Intermediate | 283 (32) |
| High | 259 (30) |
| RORP | |
| Low proliferation | 177 (20) |
| Intermediate proliferation | 485 (56) |
| High proliferation | 209 (24) |
| PAM50 | |
| Luminal A | 400 (46) |
| Luminal B | 241 (28) |
| Her2-enriched | 83 (9) |
| Basal-like | 84 (10) |
| Normal-like | 63 (7) |

Supplementary Table 2: Multivariable analysis for all the elderly patients, ER+/LN-/HER2- patients for GGI, 70-gene, RS, CCS, ROR-P, and PAM50 signatures. We used RFS as the clinical endpoint.

| | | Patients (ER+/LN-/HER2-)* (N = 222) | |
|---|---|---|---|
| *Signature* | *N(%)* | *HR (95% CI)* | *P* |
| Genomic grade index | | | |
| GG1 (ref) | 131 (59) | 1 (-) | - |
| GG3 | 91 (41) | 1.6 (0.8 - 3.3) | 0.17 |
| 70-gene | | | |
| Low risk (ref) | 149 (67) | 1 (-) | - |
| High risk | 73 (33) | 2.3 (1.2 - 4.4) | **0.02** |
| Recurrence score | | | |
| Low risk (ref) | 59 (26) | 1 (-) | - |
| Intermediate risk | 77 (35) | 0.9 (0.3 - 3.2) | 0.75 |
| High risk | 86 (39) | 1.3 (0.6 - 2.2) | 0.5 |
| Cell cycle score | | | |
| Low (ref) | 102 (46) | 1 (-) | |
| Intermediate | 76 (34) | 2.2 (1 - 4.8) | 0.05 |
| High | 44 (20) | 2.4 (1 - 5.9) | 0.06 |
| RORP | | | |
| Low proliferation (ref) | 62 (28) | 1 (-) | - |
| Int. proliferation | 130 (59) | 1.5 (0.6 - 3.6) | 0.33 |
| High proliferation | 30 (13) | 2.3 (0.7 - 7.4) | 0.15 |
| PAM50 | | | |
| Luminal A (ref) | 129 (58) | 1 (-) | - |
| Luminal B | 60 (27) | 2.3 (1.1 - 4.7) | **0.03** |
| Her2-enriched | 9 (4) | 3.2 (0.9 - 11.6) | 0.07 |
| Basal-like | 6 (3) | 0 (0 - Inf) | 1 |
| Normal-like | 18 (8) | 1.2 (0.3 - 4.3) | 0.73 |

NOTE: Bold values indicate P < 0.05.

* Adjusted for tumor size, tumor grade, and hormonal therapy.

Supplementary table 3: Additional prognostic value of gene signatures based on delta likelihood ratio ($\Delta LR$) on top of tumor grade, tumor size, and estrogen and lymph node status for all patients, and on top of tumor grade and tumor size for ER+/LN+ and ER+/LN-.

| | All patients[a] (N = 871) | | | Patients (ER+/LN+)[b] (N = 335) | | | Patients (ER+/LN-)[b] (N = 374) | | |
|---|---|---|---|---|---|---|---|---|---|
| *Signature* | $\Delta LR$ | *P* | *c-index* | $\Delta LR$ | *P* | *c-index* | $\Delta LR$ | *P* | *c-index* |
| Genomic grade index | 6.24 | **0.01** | 0.67 | 6.67 | **0.01** | 0.62 | 1.43 | 0.23 | 0.62 |
| 70 gene | 11.47 | **<0.001** | 0.68 | 5.84 | **0.01** | 0.64 | 5.88 | **0.01** | 0.65 |
| Recurrence score | 14.11 | **<0.001** | 0.68 | 13.59 | **<0.01** | 0.64 | 2.14 | 0.34 | 0.64 |
| Cell cycle | 16.97 | **<0.001** | 0.69 | 13.19 | **<0.01** | 0.65 | 6.47 | **0.04** | 0.65 |
| RORP | 22.42 | **<0.001** | 0.69 | 13.88 | **<0.001** | 0.64 | 5.49 | 0.06 | 0.64 |
| PAM50 | 11.35 | **0.02** | 0.68 | 8.02 | 0.09 | 0.62 | 8.37 | 0.08 | 0.66 |

NOTE: Bold values indicate P < 0.05.

a Adjusted for tumor size, tumor grade, estrogen receptor status, lymph node status, and hormonal therapy.

b Adjusted for tumor size, tumor grade, and hormonal therapy.

Supplementary table 4: Clinico-pathological characteristics of the ER+/LN- patients between 55 and 65 years old.

| | Patients (N = 478) | |
|---|---|---|
| | Number | Percent |
| Primary tumor characteristics | | |
| Progesterone receptor status | | |
| Positive | 219 | 23.6 |
| Negative | 113 | 45.8 |
| Unknown | 146 | 30.6 |
| HER2 | | |
| Positive | 30 | 6.3 |
| Negative | 272 | 56.9 |
| Unknown | 176 | 36.8 |
| Elston-Ellis tumor grade | | |
| *1* | 96 | 11.0 |
| *2* | 387 | 44.4 |
| *3* | 323 | 37.1 |
| Unknown | 65 | 7.5 |
| Tumor size (cm) | | |
| $<2$ | 228 | 47.7 |
| $\geq 2$ | 219 | 45.8 |
| Unknown | 31 | 6.5 |
| Treatment | | |
| Chemotherapy | 1 | 0.2 |
| Hormonotherapy | 156 | 32.6 |
| Chemotherapy + Hormonotherapy | 18 | 3.8 |
| Untreated | 257 | 53.8 |
| Unknown | 46 | 9.6 |

Supplementary Table 5: Multivariable analysis for ER+/LN- patients between 55 and 65 years for GGI, 70-gene, RS, CCS, ROR-P, and PAM50 signatures. We used RFS as the clinical endpoint.

‘

| | | Patients 55-65 (ER+/LN-)* (N = 478) | | | |
|---|---|---|---|---|---|
| *Signature* | *N(%)* | *HR (95% CI)* | *P* | $\Delta LR$ | *P* |
| Genomic grade index | | | | | |
| GG1 (ref) | 268 (56) | 1 (-) | - | 6.23 | **0.01** |
| GG3 | 210 (44) | 1.8 (1.1 - 2.8) | **0.01** | | |
| 70-gene | | | | | |
| Low risk (ref) | 297 (62) | 1 (-) | - | 10.11 | **<0.01** |
| High risk | 181 (38) | 2 (1.3 - 3.1) | **<0.01** | | |
| Recurrence score | | | | | |
| Low risk (ref) | 118 (25) | 1 (-) | - | | |
| Intermediate risk | 115 (24) | 1.4 (0.7 - 2.8) | 0.39 | 6.59 | **0.04** |
| High risk | 245 (51) | 2.1 (1.1 - 3.9) | **0.02** | | |
| Cell cycle score | | | | | |
| Low (ref) | 200 (42) | 1 (-) | | | |
| Intermediate | 158 (33) | 2.1 (1.2 - 3.7) | **<0.01** | 8.93 | **0.01** |
| High | 120 (25) | 2.1 (1.2 - 3.9) | **0.01** | | |
| RORP | | | | | |
| Low proliferation (ref) | 151 (31) | 1 (-) | - | | |
| Intermediate proliferation | 223 (47) | 2.7 (1.4 - 5.2) | **<0.01** | 12.25 | **<0.01** |
| High proliferation | 104 (22) | 2.8 (1.4 - 5.8) | **<0.01** | | |
| PAM50 | | | | | |
| Luminal A (ref) | 239 (50) | 1 (-) | - | | |
| Luminal B | 132 (28) | 1.6 (1 - 2.7) | 0.07 | | |
| Her2-enriched | 47 (10) | 1.8 (1 - 3.4) | 0.07 | 6.90 | 0.14 |
| Basal-like | 19 (4) | 2.3 (1 - 5.6) | 0.06 | | |
| Normal-like | 41 (8) | 1 (0.4 - 2.3) | 0.93 | | |

NOTE: Bold values indicate P < 0.05.

* Adjusted for tumor size, tumor grade, and hormonal therapy.

Supplementary table 6: Gene signature risk stratification of ER+/LN- elderly patients and patients between 55 and 65 years old.

| *Signature* | Patients ≥ 70 (N = 374) | Patients (55 ≤ age ≤ 65) (N = 478) | *P* |
|---|---|---|---|
| GGI | | | |
| GG1 | 234 (63) | 268 (56) | 0.06 |
| GG3 | 140 (37) | 210 (44) | |
| 70-gene | | | |
| Low risk | 249 (67) | 297 (62) | 0.20 |
| High risk | 125 (33) | 181 (38) | |
| Recurrence score | | | |
| Low risk | 106 (29) | 118 (25) | |
| Intermediate risk | 102 (27) | 115 (24) | 0.14 |
| High risk | 166 (44) | 245 (51) | |
| Cell cycle score | | | |
| Low | 166 (44) | 200 (42) | |
| Intermediate | 130 (35) | 158 (33) | 0.34 |
| High | 78 (21) | 120 (25) | |
| RORP | | | |
| Low proliferation | 108 (29) | 151 (31) | |
| Intermediate proliferation | 213 (57) | 223 (47) | **<0.01** |
| High proliferation | 53 (14) | 104 (22) | |
| PAM50 | | | |
| Luminal A | 210 (56) | 239 (50) | |
| Luminal B | 103 (28) | 132 (28) | |
| Her2-enriched | 20 (5) | 47 (10) | 0.11 |
| Basal-like | 11 (3) | 19 (4) | |
| Normal-like | 30 (8) | 41 (8) | |

NOTE: Correlations were calculated using $\chi^2$ test unless otherwise specified. Bold values indicate P < 0.05.
