## Supplementary Figures for "Clinically relevant gene signatures provide independent prognostic information in older breast cancer patients"

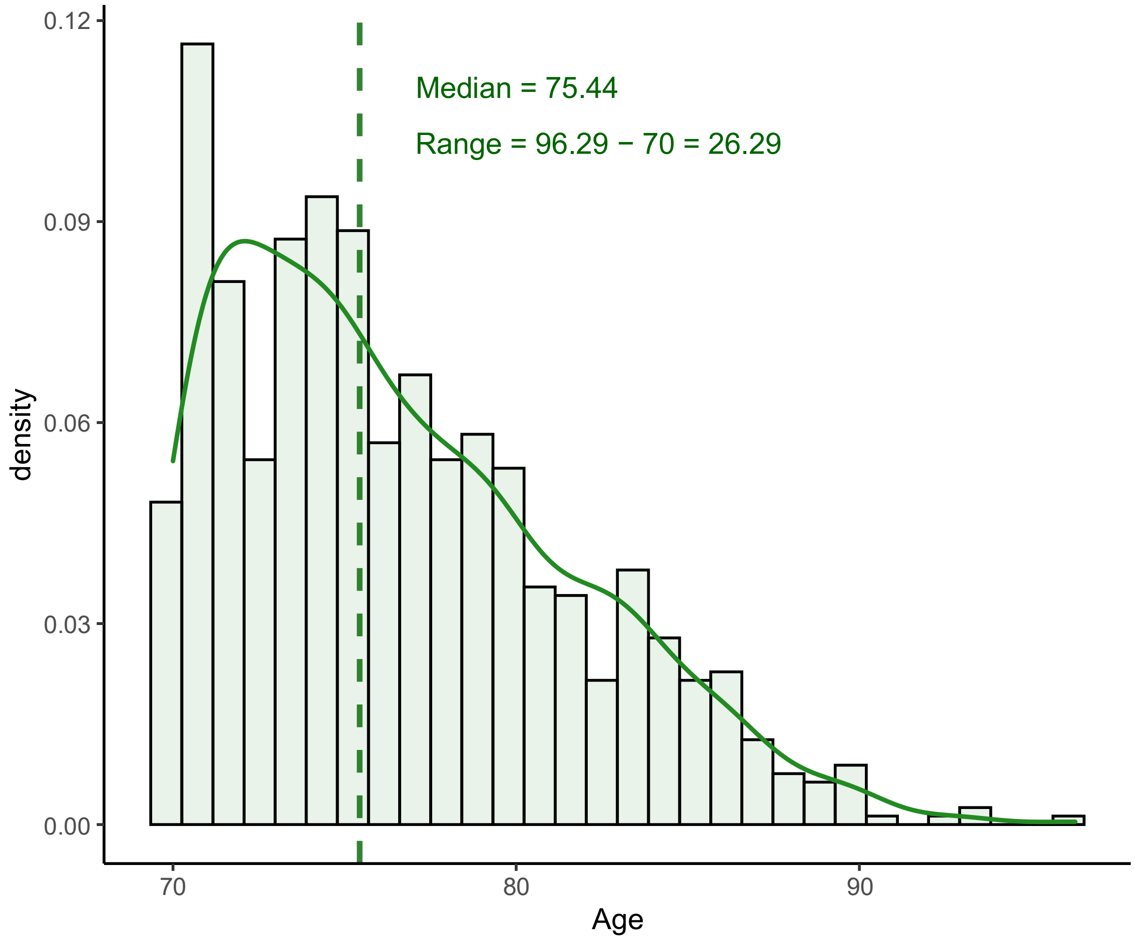


Supplementary figure 1: Age of the 871 patients from the older cohort.

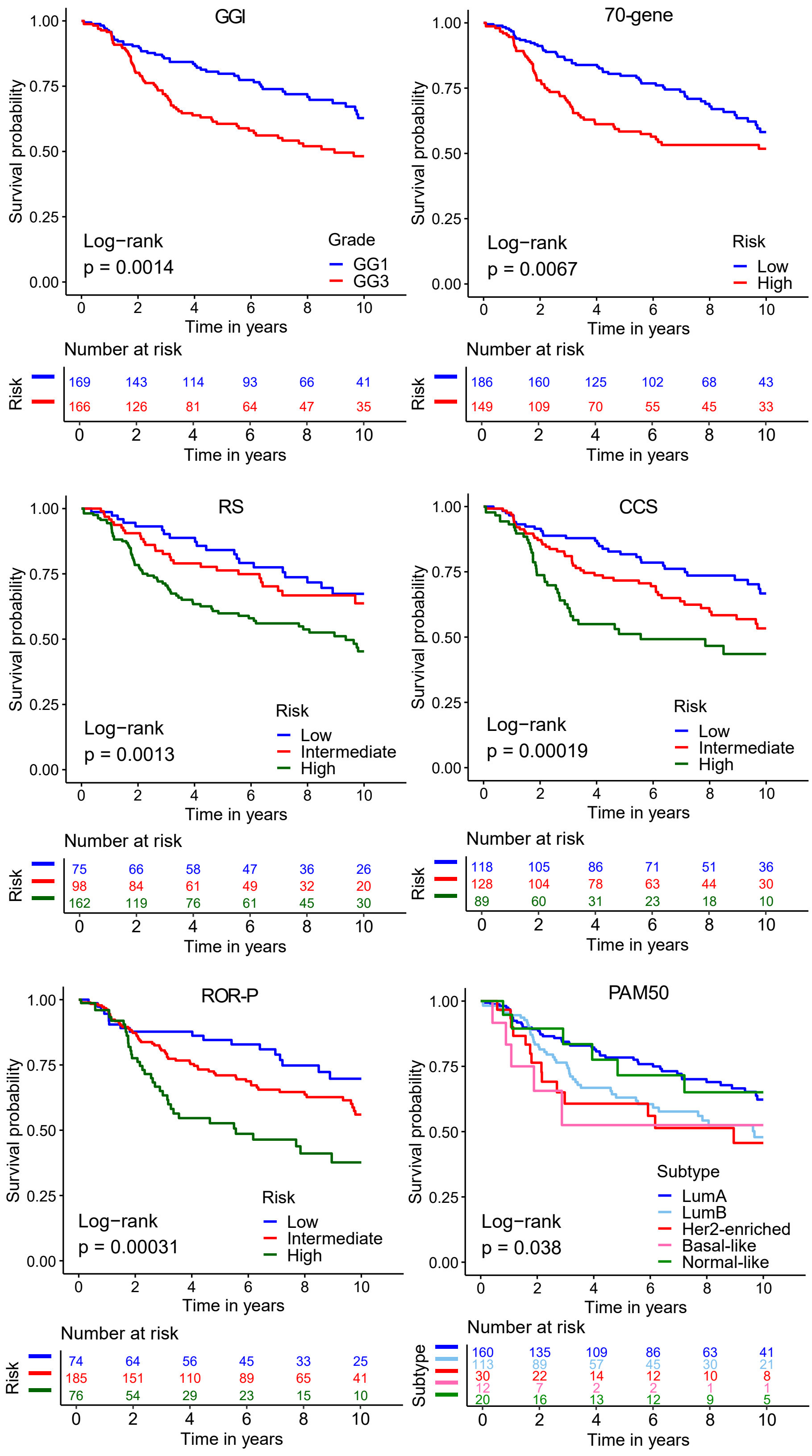


Supplementary Figure 2: Kaplan-Meier analysis of gene expression signatures in the ER+/LN+ patients. (a) Genomic Grade Index (GGI) (b) 70-gene (c) Recurrence score (RS) (d) Cell-cycle (CCS) (e) PAM50 Risk of Recurrence score - Proliferation (ROR-P) (f) Prediction Analysis of Microarray 50 (PAM50).

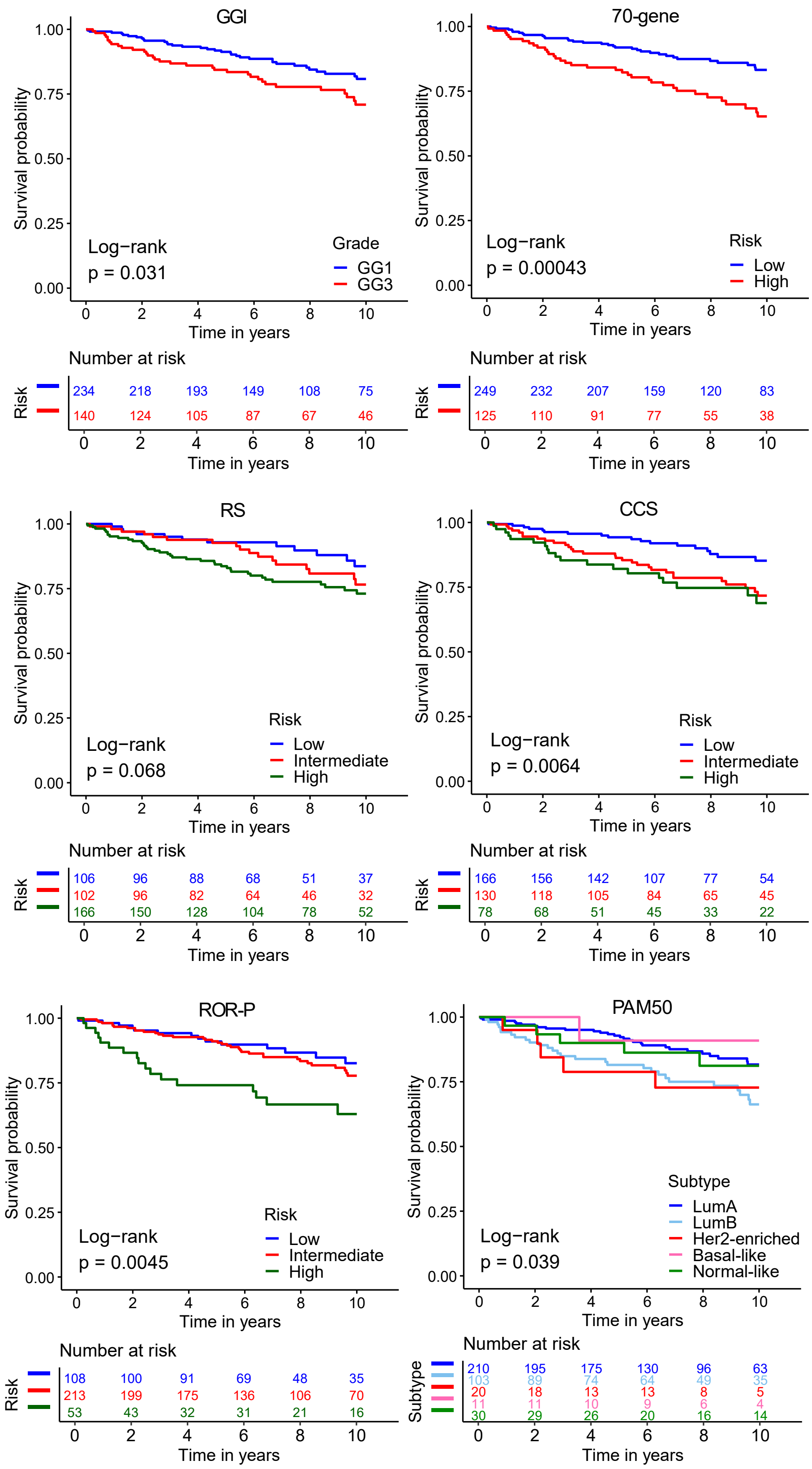


Supplementary Figure 3: Kaplan-Meier analysis of gene expression signatures in the ER+/LN- patients. (a) Genomic Grade Index (GGI) (b) 70-gene (c) Recurrence score (RS) (d) Cell-cycle (CCS) (e) PAM50 Risk of Recurrence score - Proliferation (ROR-P) (f) Prediction Analysis of Microarray 50 (PAM50).
